# Inflammatory reprogramming of the tumor microenvironment by infiltrating clonal hematopoiesis is associated with adverse outcomes in solid cancer

**DOI:** 10.1101/2024.08.29.24312110

**Authors:** Marco M. Buttigieg, Caitlyn Vlasschaert, Alexander G. Bick, Robert J. Vanner, Michael J. Rauh

## Abstract

Clonal hematopoiesis (CH) – the expansion of somatically-mutated hematopoietic cells in blood – is common in solid cancers. CH is associated with systemic inflammation that may lead to cancer, but its impact on tumor biology is underexplored. Here, we report the effects of CH on the tumor microenvironment (TME) using 1,550 treatment-naïve patient samples from the CPTAC cohort. CH was present in 18.3% of patients, with one-third of CH mutations also detectable in tumor-derived DNA from the same individual (CH-Tum), reflecting CH-mutant leukocyte infiltration. The presence of CH-Tum was associated with worse survival across cancers, particularly for glioblastoma.

Transcriptomics and proteomics revealed that CH drives inflammation in the TME in a cancer- and CH driver-specific manner, and may improve immunotherapy responses. In glioblastoma, CH associated with pronounced macrophage infiltration, inflammation, and an aggressive, mesenchymal phenotype. Our findings demonstrate that CH shapes the TME, with potential applications as a biomarker in precision oncology.

## Introduction

CH describes the clonal expansion of somatically mutated hematopoietic stem cells (HSCs) with a fitness advantage.^1–3^ CH is detected in blood of approximately 20-30% of patients with solid cancers, which is considerably higher than the reported prevalence in comparably-aged cancer-free individuals.^4,5^ CH driver mutations, including inactivation of the most common driver genes *DNMT3A* or *TET2*, are associated with systemic inflammation.^1,6,7^ Accordingly, CH is associated with excess morbidity and mortality attributable to numerous chronic diseases;^1,8–12^ however, its impact on solid tumors remains unclear.^1,8–12^

The higher prevalence of CH in solid cancer patients relative to the general population is related to age, exposure to cancer therapy, and shared risk factors like smoking or germline genetics.^4,5^ Consequently, there is growing interest in how CH effects the tumor microenvironment (TME) and influences cancer outcomes.^13^ CH has been associated with increased risk of some solid cancers in several cohorts.^14–20^ The mechanisms for this remain unclear, but it is possible that CH-related inflammation promotes tumorigenesis in certain disease contexts like lung and liver cancers.^19,20^ Furthermore, associations between CH and survival are heterogeneous and context-dependent, with favourable effects reported in metastatic colorectal cancer patients, no effect in some single-centre cohorts, and negative effects in pan-cancer cohorts subject to confounding by disease stage and treatment history.^4,18,21–24^ While CH in the human TME may influence inflammation in advanced tumors,^25^ the evidence is limited and clinical relevance remains uncertain.

Herein, we present a pan-cancer study of CH as a feature of solid tumor biology, studying 1,550 primary, treatment naïve patients from the Clinical Proteomics Tumor Analysis Consortium (CPTAC). By integrating genomic, transcriptomic, proteomic, and clinical data, we identify the presence of CH mutations in tumor sequencing (CH-Tum) as a critical determinant of the effects of CH in solid cancer, highlighting its potential as a biomarker for precision oncology.

## Results

### Clonal hematopoiesis mutations are commonly detected in the blood and tumors of treatment-naïve cancer patients

To identify patients in the CPTAC cohort with CH, we re-analyzed peripheral blood whole exome sequencing (WES) data using our established protocol described by Vlasschaert *et al*.^26^ The cancers represented in CPTAC (n=1,550; Table S1) include uterine corpus endometrial carcinoma (UCEC; n=240), lung adenocarcinoma (LUAD; n=229), clear cell renal cell carcinoma (ccRCC; n=214), pancreatic ductal adenocarcinoma (PDAC; n=163), breast cancer (BRCA; n=130), head and neck squamous cell carcinoma (HNSC; n=110), lung squamous cell carcinoma (LUSC; n=108), colorectal adenocarcinoma (COAD; n=106), glioblastoma multiforme (GBM; n=104), high grade serous ovarian cancer (OV; n=90), and fewer samples from various sites (Other; n=56). All patients were treatment naïve at the time of sample collection, with primary tumor samples collected from both local (stage I-III, n=1,263) and metastatic (stage IV, n=218) cancers.

Defining CH, also known as CH of indeterminate potential (CHIP), as the presence of a somatic driver mutation in peripheral blood WES with variant allele fraction (VAF) ≥2% (Figure 1A), the prevalence of CH in the CPTAC cohort was 18.3% (n=283; Figure 1B). CH was significantly associated with patient age across the CPTAC cohort (p=1.6×10^-9^, rank-sum test; Figures 1C, S1A-B), and in all cancers except COAD (p=0.69, rank-sum test). There were significant variations in the CH prevalence across different cancer types in CPTAC, even after accounting for patient age and sex. Compared to the cohort average weighted by cancer type, OV and COAD had higher rates of CH (OV odds ratio [OR]=2.35 [1.41-3.91], COAD OR=1.61 [1.04-2.48], binomial regression). CH was not associated with more advanced tumor stage at the time of diagnosis (OR=1.26 [0.98-1.62], ordered logistic regression; Figure S1C). The mutational spectrum of CH was consistent with previous reports (Figures 1D-E; Table S2). Mutations in *DNMT3A* (37.9% of CH; n=132) and *TET2* (20.7%; n=72) composed the majority of CH drivers, with VAF ranging from 0.02-0.38 (median VAF=0.05). Most patients presented with only one CH mutation (81.3%; n=230), but 53 patients (18.7%) had between 2-4 CH drivers (Figure S1D).

**Figure 1:**
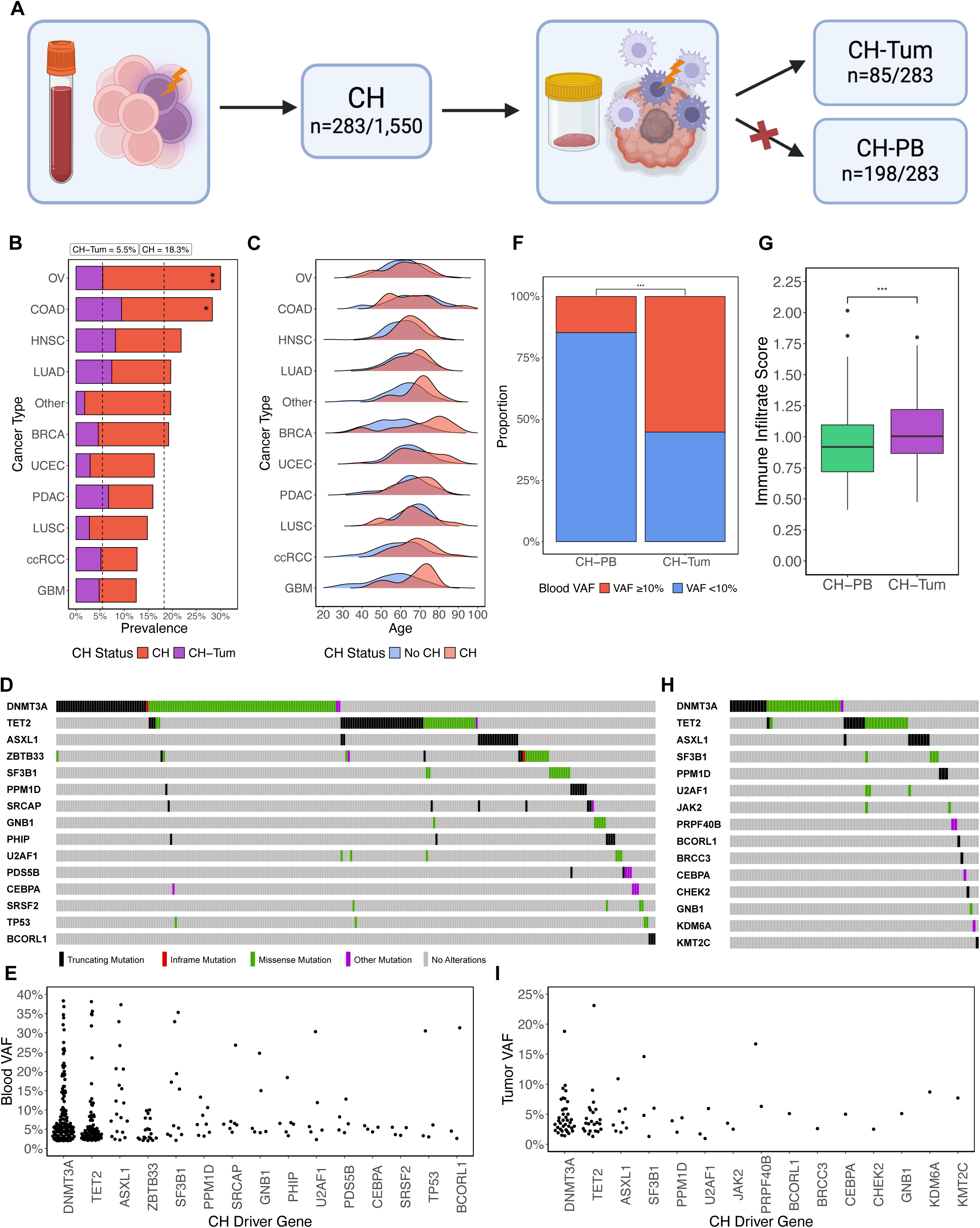
Survey of the characteristics of CH and CH-Tum in treatment-naïve solid cancer patients. (A) Schematic overview of CH calling and definitions of CH-Tum and CH-PB. Any patient with CH driver mutations in peripheral blood WES was classified as having CH. Of those with CH, if the same mutations were subsequently detected in tumor WES, they were labelled as CH-Tum. If the peripheral blood mutations were not subsequently detected in the tumor sample, they were labelled CH-PB. (B) Prevalence of CH and CH-Tum by cancer type. (C) Age distributions of patients with and without CH by cancer type. (D) Oncoplot depicting the mutational spectrum of CH. (E) Peripheral blood VAFs of CH mutations by driver gene. (F) Proportions of large (VAF ≥10%) and small (VAF 2-10%) CH clones that make up CH-PB and CH-Tum. (G) Immune infiltrate scoring by CIBERSORTx in CH-Tum and CH-PB. (H) Oncoplot depicting the mutational spectrum of CH-Tum. (I) Tumor VAFs of CH-Tum mutations by driver gene.

CH mutations are detectable in tumor-derived DNA sequencing.^4,5,27^ Since CH mutations are of hematopoietic origin, we posited that their identification in tumor sequencing reflects a greater degree of infiltration by CH-mutant leukocytes into the TME, potentially identifying cases where CH most potently impacts tumor biology. After identifying CH in blood sequencing, we defined the detection of the same CH driver mutations in tumor sequencing as CH-Tum (Figure 1A; STAR Methods). The prevalence of CH-Tum was 5.5% (n=85), accounting for 30% of patients with CH (Figure 1B).

Mutations in the remaining 70% of patients with CH solely found in peripheral blood are referred to as CH-PB (Figure 1A). Like CH, CH-Tum increased significantly with patient age (p= 7.9×10^-5^, rank-sum test; Figure S1B). Amongst those with CH, the presence of CH-Tum did not vary by cancer type after accounting for age (Figure 1B). In a multivariate model we found that detection of CH-Tum was associated with CH mutation VAF ≥10% in the peripheral blood (OR=9.21 [4.58-18.6], binomial regression; Figure 1F) and degree of immune infiltration in the tumor sample as quantified by CIBERSORTx (OR=6.69 [1.96-22.9], binomial regression; Figure 1G). Immune infiltrate scoring did not significantly differ based on peripheral blood VAF in CH-Tum or CH-PB (CH-Tum p=0.19, CH-PB p=0.2, rank-sum test; Figure S1E), but when separating between VAF ≥10% and <10%, immune infiltrate was significantly higher in CH-Tum relative to CH-PB in both conditions (VAF≥10% p=0.02, VAF<10% p=0.002, rank-sum test; Figure S1F).

This suggests that CH-Tum captures both tumor-immune infiltration and CH clone size, rather than acting solely as a surrogate for large CH clones in the blood. Similar to CH, CH-Tum was not associated with more advanced tumor stage at the time of diagnosis (OR=1.41 [0.94-2.13], ordered logistic regression; Figure S1G).

The distribution of driver mutations in CH-Tum closely resembled that of CH, with *DNMT3A* (40% of CH-Tum, n=38) and *TET2* (24.2%, n=23) predominating (Figures 1H-I; Table S2). There were no discernable patterns of enrichment of any prominent driver genes in CH-Tum compared to CH. Mutation VAF in CH-Tum ranged from 0.01-0.23 and was positively correlated with peripheral blood VAF (median tumor VAF=0.04, median blood VAF=0.12; Spearman’s rho=0.54, p=1.9×10^-8^, Spearman’s rank correlation; Figure S1H).

We also evaluated 8,927 matched samples collected for The Cancer Genome Atlas (TCGA) project (n=8,927; Table S1) from 32 cancer types to identify CH and CH-Tum. CH driver mutations were found in the peripheral blood WES of 780 individuals, yielding a CH prevalence of 8.74% (median VAF=0.067; Table S2). CH-Tum was also detectable in TCGA tumor samples. Of the 780 TCGA participants with CH, we were able to subsequently identify at least one of their CH driver mutations in the tumor WES for 122 (15.6%), representing 1.3% of the entire cohort (median tumor VAF=0.05, median blood VAF=0.17; Table S2).

The prevalence of CH and CH-Tum in the TCGA were lower than the CPTAC cohort and prior cancer cohorts profiled with targeted-sequencing.^4,5,27^ Comparing CPTAC and TCGA directly, patients in CPTAC were significantly more likely to have CH, *DNMT3A*-CH and *TET2*-CH, even after accounting for differences in patient age and sex (CH: OR=1.93 [1.63-2.27]; *DNMT3A*-CH: OR=2.45 [1.92-3.12]; *TET2*-CH: OR=3.74 [2.60-5.38], binomial regression; Figures S1I-K).

Given the significant gaps in coverage related to the exome capture kits used by the TCGA sequencing sites,^28–30^ we compared the depth of coverage in CPTAC and TCGA peripheral blood WES samples (Table S3). Across coding regions of CH driver genes considered in this study, CPTAC samples had a median average depth of 259x, which was significantly higher than the average depth in TCGA samples of 94x (p=0, rank-sum test; Figure S1L). This phenomenon was present for *DNMT3A* (CPTAC median coverage=215x, TCGA median coverage=77x; p=0, rank-sum test; Figure S1M) and even more pronounced for *TET2* (CPTAC median coverage=312x, TCGA median coverage=45x; p=0, rank-sum test; Figure S1N), with a number of TCGA samples receiving no coverage at all in these critical CH driver genes. Furthermore, although coverage depth did not differ based on CH status in CPTAC, inconsistencies in coverage depth within the TCGA cohort led to the average depth being significantly higher in samples identified to have CH (CPTAC: p=0.45; TCGA: p=2×10^-17^, rank-sum test; Figures S1O-P). Noting these limitations, we largely excluded this cohort from subsequent analyses.

### CH-mutant leukocyte infiltration of the TME is associated with adverse clinical outcomes

In Cox proportional hazard regression analyses adjusted for age, sex, cancer type, smoking status, and disease stage (hereafter, “adjusted”), CH status was not associated with overall survival in the CPTAC cohort (univariate hazard ratio [HR]=1.07 [0.82-1.40], multivariate HR=1.12 [0.83-1.50]; Figures 2A-B) or within specific cancer sub-cohorts (Figures 2B, S2A). However, the presence of CH-Tum was associated with worse overall survival across the CPTAC cohort (univariate HR=1.61 [1.07-2.42], multivariate HR=1.74 [1.13-2.69]; Figures 2C-E). Despite statistical power limitations for analyses stratified by cancer type, CH-Tum was associated with poor survival in GBM (univariate HR=3.60 [1.39-9.29], multivariate HR=3.63 [1.21-10.9]) and trended towards worse survival in PDAC (univariate HR=1.94 [0.93-4.05], multivariate HR=2.11 [1.00-4.46]; Figures 2D, S2B). Unlike CH-Tum, immune infiltration was not independently associated with overall survival (univariate HR=1.29 [0.88-1.90]; Figure S2C), nor did its inclusion in multivariate analysis negate the survival association of CH-Tum (CH-Tum HR=1.83 [1.18-2.84]). This suggests that the effects of CH on survival are specifically related to CH-Tum and not simply elevated immune infiltration. To help elucidate the root of this excess mortality, the last known tumor status at follow up was modelled for CH and CH-Tum in adjusted survival analysis. CH-Tum, but not CH, was associated with a reduced likelihood of being classified as tumor-free at follow up (CH OR=0.67 [0.43-1.08], CH-Tum OR=0.38 [0.17-0.81]; Figure 2F).

**Figure 2:**
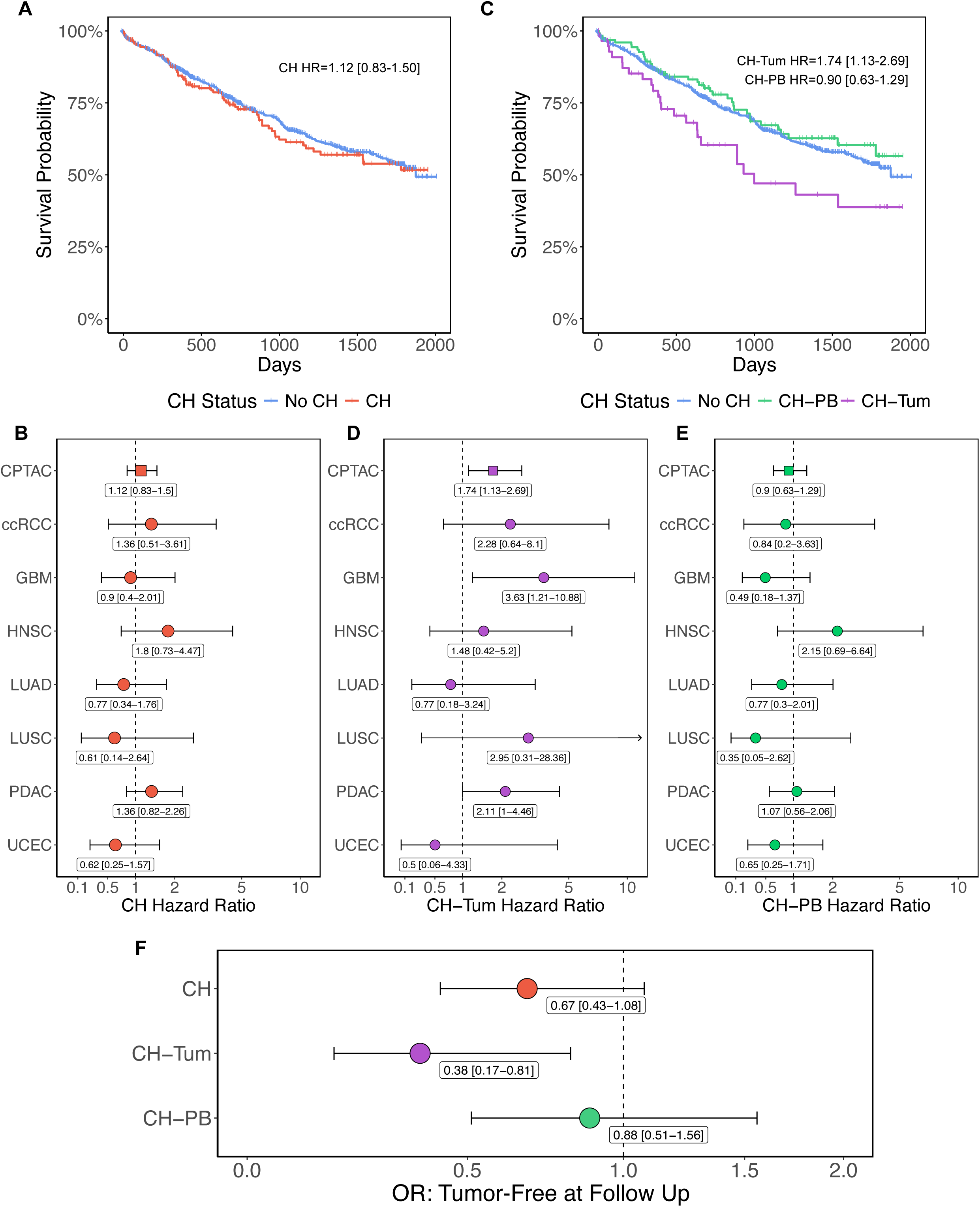
Clinical associations of CH and CH-Tum in solid cancer patients. (A) Kaplan-Meier plot comparing overall survival in those with and without CH. (B) Forest plot and hazard ratios from multivariate cox proportional hazard regression for overall survival by CH status. (C) Kaplan-Meier plot comparing overall survival in those with CH-Tum, CH-PB, and no CH. (D-E) Forest plot and hazard ratios from multivariate cox proportional hazard regression for overall survival by CH-Tum (D) and CH-PB (E). (F) Forest plot and odds ratios from multivariate binomial regression of likelihood of a patient being classified as tumor-free at last known follow up.

### Clonal hematopoiesis is associated with an inflamed tumor microenvironment

Next, we explored the impact of CH on tumor biology using bulk tumor RNA-sequencing and proteomics. Compared to tumors in patients without CH, there were 363 and 301 differentially expressed genes with CH and CH-Tum, respectively, after accounting for sex and cancer type (Figures 3A-B). In both CH and CH-Tum, we observed the upregulation of genes related to immunity and tumor-immune interactions including *SPINK1*,^31–33^ cytokines *IL1A*, *IL17REL*, and *CXCL6*, and S100 alarmins. Of these differentially expressed genes, 96 were shared across CH and CH-Tum positive tumors compared to no CH, with the 64 shared upregulated genes exhibiting significantly greater log fold changes in the CH-Tum condition (p=8.5×10^-9^, pairwise rank-sum test; Figure 3C). Despite limitations with CH detection in the TCGA cohort, multiple differentially expressed genes in CH and CH-Tum from CPTAC were also found in CH-Tum positive cases from the TCGA (Figures S3A-B). 54 genes were similarly dysregulated with CH-Tum in CPTAC and TCGA, including upregulation of *CTLA4*, *LILRA2*/*A5*/*B2*, *SPINK1*, and *TLR2*/*8*. Functional profiling of these shared upregulated genes in CH-Tum showed enrichment of pathways related to antigen presentation and IL-6 signalling (Figure S3C).

**Figure 3:**
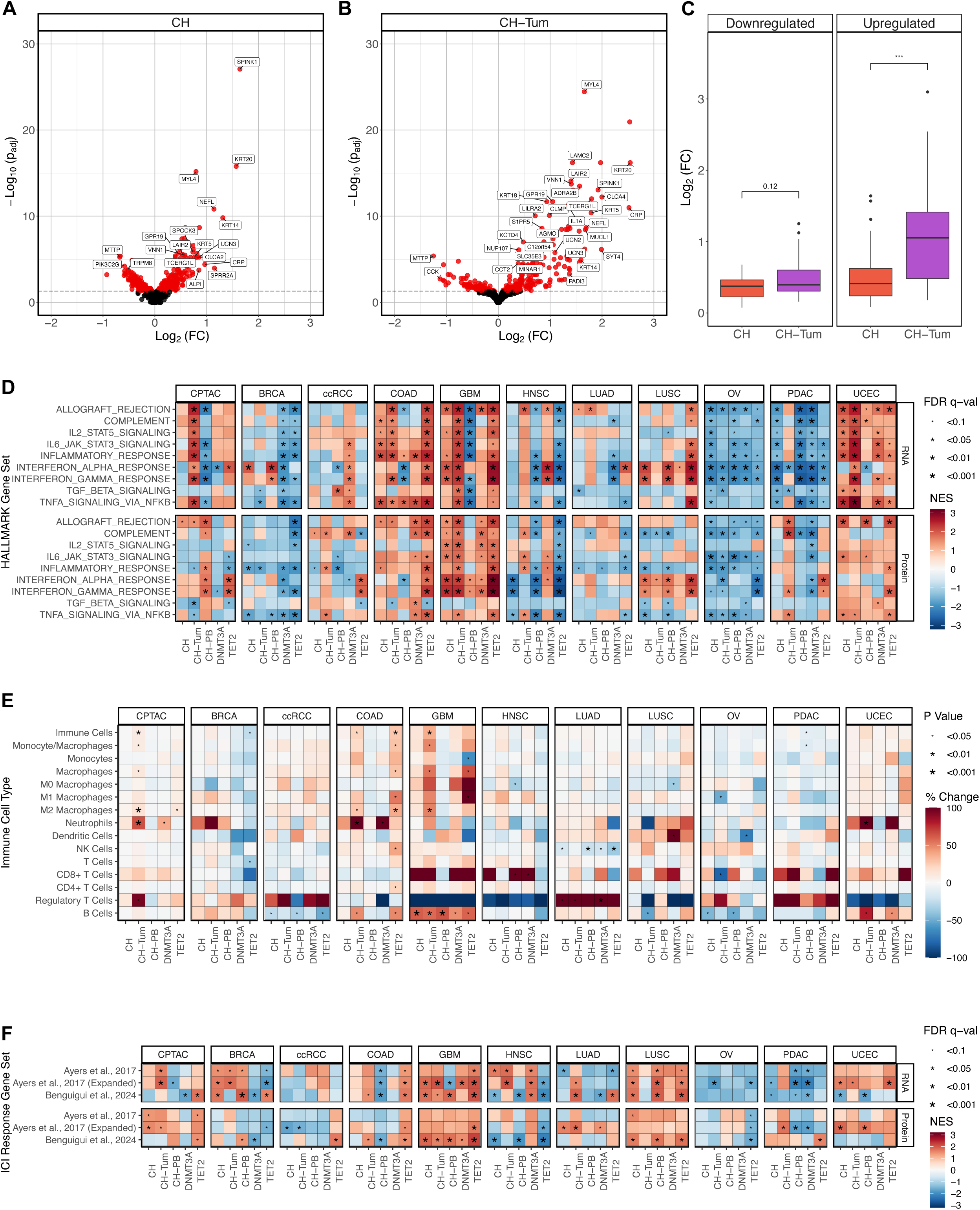
Transcriptomic and proteomic associations of CH in the TME. (A-B) Volcano plots depicting differential tumor gene expression in CH (A) and CH-Tum (B) versus no CH across the CPTAC pan-cancer cohort. (C) Log fold changes of shared differentially expressed genes in CH versus CH-Tum. (D) RNA and protein GSEA of select immune-related HALLMARK gene sets by CH status and cancer type. (E) Enrichment of select LM22 immune cell subpopulations in the tumor by CH status and cancer type. Cells with percent change ≥+100% are colored as +100% to preserve data visualization. (F) RNA and protein GSEA of anti-PD-1 ICI predicted response gene sets by CH status and cancer type.

We then performed gene set enrichment analysis (GSEA) using differential gene expression and mass-spectrometry-based protein abundance analysis of tumor samples in the CPTAC dataset. CH and CH-Tum, as well as CH with *DNMT3A* and *TET2* driver mutations, were associated with marked changes in HALLMARK gene sets related to inflammation and immunity (Figure 3D; Table S4). Across all cancers in CPTAC and controlled for cancer type, CH-Tum was associated with transcriptomic enrichment of immune signatures in tumors compared to patients without CH (Figure 3D). Contrarily, CH-PB was associated with decreased inflammatory gene expression across cancers at the transcriptomic level (Figure 3D). While most individual cancer types had agreement between RNA and protein modalities, there was limited concordance at the pan-cancer level that may be related to sampling bias within the UCEC cohort, where fewer than half of tumors profiled by RNA-seq (n=223) also underwent proteomic analysis (n=97). A pan-cancer signal consistent across transcriptomic and proteomic profiling was enrichment of the interferon (IFN) response with *TET2*-driven CH. This was consistent across cancer types, where IFN signalling pathways were frequently dysregulated (Figure 3D). In addition to immune-related signalling, CH also associated with enrichment of canonical cancer pathways including angiogenesis, metabolism, and oncogenic signalling that may drive tumor growth and progression, underlying adverse clinical associations (Figure S3D). There was notable heterogeneity between CH-associated immune signatures across cancer types. Cancers like COAD, GBM, and UCEC were marked by a pro-inflammatory phenotype found with both CH and CH-Tum, while BRCA, OV, and PDAC exhibited reduced inflammation (Figure 3D). CH driven by mutations in *TET2* displayed a more prominent effect than *DNMT3A* among cancers with a broad pro-inflammatory signature such as COAD, GBM, and LUSC (Figure 3D).

To evaluate differences in immune cell composition in the TME, we compared CIBERSORTx-quantified cell fractions in those with and without CH, again adjusted for cancer type, patient sex, and other cancer-specific factors using linear regression (Figure 3E). There was a 7.67% increase in overall immune infiltrate in CH-Tum across all cancers (p=0.008), along with prominent signatures of elevated immune infiltration in COAD and GBM seen with CH-Tum (COAD: +22.6%, p=0.02; GBM: +36.7%, p=0.004; Figure 3E). This enrichment was largely due to increases in the abundance of common myeloid cells including macrophages (+10.3%, p=0.01) and neutrophils (+65.1%, p=9×10^-4^; Figure 3E). Although regulatory T cells make up a small fraction of the total immune cell population, they were also found to be enriched across cancers with CH-Tum (+95.4%, p=0.04; Figure 3E). Despite the association between CH and immune infiltration, the observed cancer-specific effects of CH were independent of the degree of immune infiltrate seen with each cancer studied; the effects of CH were not isolated to tumor sites classified as immunologically ‘hot’ or ‘cold’ (Figure S3E). CH and CH-Tum status were also independent of broad, proteogenomics-defined tumor immunological subtypes (Figure S3F).^34^

Since CH has been associated with survival in patients treated with ICI,^22^ we tested for enrichment of three pan-cancer validated gene signatures predictive of anti-PD-1 ICI response.^35,36^ CH-Tum and *TET2*-driven CH both showed significant enrichment of ICI response signatures at the transcriptomic and proteomic level (Figure 3F; Table S4). *TET2*-driven CH was particularly associated with predicted ICI response in COAD, GBM, and LUSC. In patients with GBM, CH and CH-Tum, but not CH-PB, were also associated with positive predicted responses (Figure 3F).

### Clonal hematopoiesis shapes glioblastoma phenotype and immune infiltration

Cancers span transcriptionally and biologically distinct molecular subtypes. We identified an association between CH and tumor molecular subtype in COAD (CMS3 OR=3.27 [1.16-9.22]; Figures S4A-H) and a strong trend towards CH-Tum being associated with the aggressive, immune-rich, mesenchymal GBM subtype^37^ (OR=6.55 [0.70-61.1], binomial regression; Figure 4A). In addition to the trend towards more mesenchymal tumors, we also noted an enrichment of glioma stem cell (GSC) marker genes^38^ via GSEA in CH-Tum in GBM (Figure 4B).

**Figure 4:**
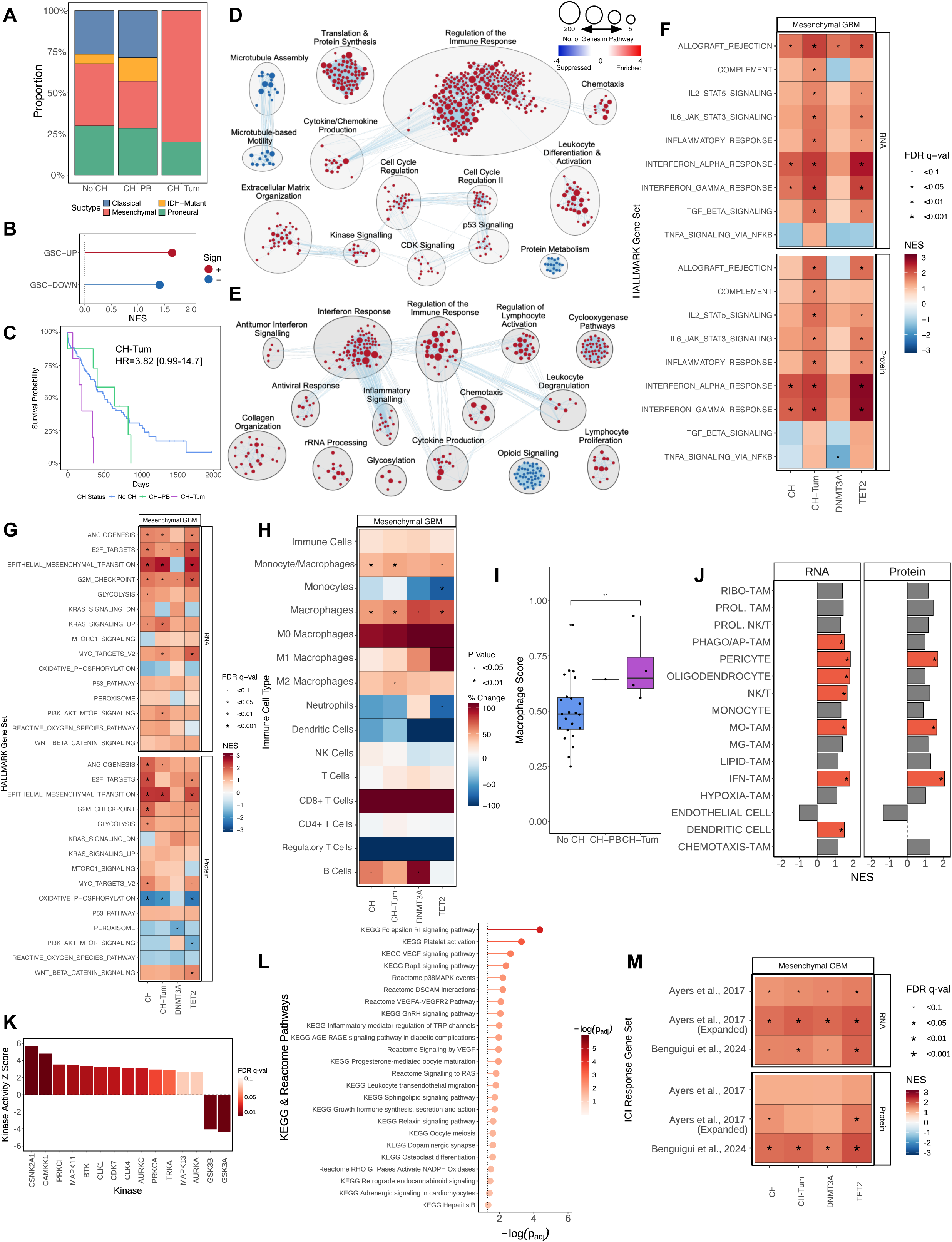
Clinical and multi-omics associations of CH in mesenchymal GBM. (A) Proportion of GBM molecular subtypes among CH-Tum, CH-PB, and no CH. (B) Lollipop plot of GSC gene signature GSEA in GBM with CH-Tum. (C) Kaplan-Meier plot comparing overall survival in mesenchymal GBM patients with CH-Tum, CH-PB, and no CH. (D-E) Cytoscape network diagrams depicting RNA (D) and protein (E) GSEA results of EnrichmentMap gene sets for CH-Tum in mesenchymal GBM. Significantly enriched (FDR<0.1) gene sets in the 15 largest clusters are displayed. Gene set nodes were clustered based on similarity, with edges representing higher set overlap. Gene sets colored red are upregulated, while those colored blue are downregulated. (F-G) RNA and protein GSEA of select immune-associated (F) and tumor-associated (G) HALLMARK gene sets by CH status in mesenchymal GBM. (H) Enrichment of select LM22 immune cell subpopulations in the tumor by CH status in mesenchymal GBM. (I) LM22 macrophage scoring in CH-Tum, CH-PB, and no CH for mesenchymal GBM. (J) RNA and protein GSEA enrichment for GBM immune cell phenotypes for CH-Tum in mesenchymal GBM. (K) KSEA significantly altered kinase activity for CH-Tum in mesenchymal GBM. (L) Significantly enriched KEGG and Reactome gene sets based on functional profiling of enriched kinases with CH-Tum in mesenchymal GBM. (M) RNA and protein GSEA of anti-PD-1 ICI predicted response gene sets by CH status in mesenchymal GBM.

Given the prominent survival association and inflammatory dysregulation seen with CH in GBM, as well as the immune involvement of mesenchymal GBM tumors, we used this cancer as a case study to further explore how CH-mutant leukocytes may alter the TME and shape tumorigenesis, even within a single cancer molecular subtype. As such, we re-evaluated survival and inflammatory associations of CH in the context of mesenchymal GBM. Cox proportional hazard regressions of CH status demonstrated that CH-Tum retained a significant effect on survival in mesenchymal GBM using a univariate model, and trends toward significance in adjusted analysis despite underpowering in this small sample subset (univariate HR=3.75 [1.17-12.1], multivariate HR=3.82 [0.99-14.7]; Figure 4C).

Transcriptome and proteome GSEA of the HALLMARK pathways revealed that in mesenchymal GBM, CH, but especially CH-Tum and *TET2*-driven CH, associate with a pro-inflammatory phenotype with enrichment of epithelial-mesenchymal transition and angiogenesis pathways (Figures 4D-G; Table S4). Differential expression analyses were controlled for *NF1* mutation status, which is known to exacerbate inflammation and immune cell recruitment in GBM.^37^ Similarly, CIBERSORTx showed that CH was associated with prominent increases in macrophage abundance in the TME (+47.4%, p=0.009) ; Figures 4H-I). Using previously derived GBM immune cell phenotypes,^39^ CH-Tum was found to be enriched for monocyte-derived tumor-associated macrophages (MO-TAMs), IFN-stimulated TAMs (IFN-TAMs), and pericytes among others (Figure 4J; Table S4). The exacerbated mesenchymal phenotype of GBM with CH-Tum was also seen at the phosphoproteomic level, with overactivity of tumor-promoting kinases MAPK11/13, PRKCA/I, and others contributing to enrichment of immune migration and pro-angiogenic signalling signatures from KEGG and Reactome pathways (Figures 4K-L). Finally, as in the GBM-wide analysis, the mesenchymal subtype samples showed enrichment of gene signatures predictive of ICI response when CH was present (Figure 4M). These data suggest that within a seemingly uniform GBM molecular subtype, CH-mutant leukocytes in the TME influence immune infiltration and inflammatory programs to shape an aggressive tumor phenotype.

## Discussion

Here we present a pan-cancer multi-omics analysis of CH in solid tumors as a resource to accelerate the development of CH as a biomarker in precision immune-oncology. We extend limited previous studies of CH in solid tumors by defining a clinically-relevant CH classification – CH-Tum – representing the infiltration of CH-mutant leukocytes into the tumor and their detection via tumor sequencing. With CH-Tum, we demonstrate that CH can influence hallmark cancer properties of immune infiltration, inflammation, angiogenesis, and mitogenic signalling. Critically, we show that clinical outcomes and gene expression changes are proportional to the degree of CH-mutant leukocyte infiltration as detected in tumour sequencing. A prior study exploring CH as a proponent of tumor-associated inflammation was limited by the use of samples from advanced cancers exposed to prior treatment, detection of CH from cell-free DNA, and limited consideration of the heterogeneity of responses between CH drivers and cancer types.^25^ By leveraging our validated CH identification pipelines in the context of the rich clinical and proteogenomic resources provided by CPTAC, we show that CH has potential as a biomarker that captures multiple molecular characteristics of a tumor, with predictive and prognostic implications.

CH-Tum has been previously identified in tumor sequencing reads as a potential contaminant confounding molecular tumor diagnostics.^4,5,27^ We demonstrate that rather than simply acting as diagnostic interference, the presence of CH-Tum is clinically and biologically significant in solid cancers, associating with worsened survival outcomes and reduced likelihood of being tumor-free following treatment. Detection of CH-Tum is feasible for clinical implementation with minor modification of existing paired tumor-blood sequencing practices. With WES at ∼250x depth, approximately one third of those with CH and 5.5% of all patients demonstrate CH-Tum, similar to previous estimates.^27^ Although presented as a binary variable in this study due to technical limitations of WES, CH-Tum (versus CH-PB) likely exists on a spectrum, and future work will be required to delineate appropriate VAF thresholds that define the most clinically relevant levels of CH-mutant leukocyte infiltration in tumors. With CH mutations recently identified in atherosclerotic lesions,^40,41^ the relevance of tissue invasion by CH-mutant leukocytes may also extend beyond solid cancers, presenting a new paradigm for understanding CH-related pathophysiology.

Through the characterization of differential gene expression and protein abundance in tumors with and without CH, we find CH and especially CH-Tum to be associated with dysregulated inflammation, tumor-intrinsic signalling, and immune infiltration in the TME. Myeloid differentiation bias of mutant HSCs and the production of functionally altered myeloid cells occurs with many CH driver mutations, including *TET2*,^42^ and may underlie the elevated presence of macrophages and neutrophils in the TME with CH-Tum. The presence of CH-mutant leukocytes in the TME may also facilitate further recruitment and infiltration of immune cells. CH-mutant leukocytes have potential to alter tumor biology and the tumor-immune interface through excessive release of pro-inflammatory cytokines that interact with wild-type immune cells, tumor cells, and stroma.^6,7,43,44^ The functional enrichment of IL-6 signalling and antigen presentation seen with CH-Tum across both CPTAC and the TCGA implicates inflammation and myeloid-lymphoid crosstalk as potential mechanisms underlying the observed changes. Additionally, increased neutrophil abundance and S100 alarmin expression suggest a contribution from tumor-associated neutrophils. Cancer-specific variation in these inflammatory interactions may underlie the heterogeneous effects of CH observed in different cancer types. It is possible that CH-mutant leukocytes consistently secrete pro-inflammatory factors, but the stromal, vascular, wild type leukocyte, and cancer cells’ responses determine outcomes.

Our results demonstrate the potential for CH, or CH-Tum, as a biomarker that may identify patients with distinct, inflammatory tumor biology and worse prognosis.

Furthermore, CH or a variant thereof, may have predictive potential if it identifies individuals whose tumors are primed for immunotherapy response. Alternatively, given the association between CH and incident inflammation-related cancers, CH may identify patients for anti-inflammatory prophylaxis. The impact of CH and potential for biomarker development is highlighted in GBM, where CH-Tum is linked to inflamed, mesenchymal tumors. Even within mesenchymal subgroup tumors, CH seems to amplify this phenotype, and is linked to immune infiltration, mitogenic, angiogenic, and stemness pathways associated with poorer outcomes. GBM is uniquely poised to be influenced by CH since mesenchymal glioma cell differentiation is induced by macrophages in the TME.^37,45,46^ CH is associated with an inflamed endothelium,^47,48^ and the observed associations between CH, angiogenesis, and elevated pericytes may reflect breakdown of the blood-brain barrier in GBM, aiding the recruitment of CH-mutant myeloid populations into tumors to elicit the mesenchymal transformation.^49,50^ Curiously, despite driving a more aggressive tumor phenotype, mesenchymal-like glioma cells may be more susceptible to cytotoxic T cells induced by anti-PD-1 ICI therapy. This may explain the apparent discrepancy between our finding of predicted ICI response with CH, but the observed worse survival in GBM patients with CH by Hsiehchen *et al*. Both studies identify poor prognostic associations of CH in GBM; however, whether mesenchymal GBM patients with CH could benefit from immunotherapy more than conventional therapy remains unknown.

Limitations of our study include tumor sample purity and sampling bias influencing CH-Tum detection in tumor sequencing, and sample size limitations for cancer- or gene-specific analyses. The lack of single-cell analysis precludes identification of the source of dysregulated gene expression pathways in patients with CH. As large-scale cancer datasets are established in the future, the availability of this data will allow for more thorough examinations of CH and its role in the TME.

CH has considerable potential as a biomarker in precision oncology beyond its known connection to therapy-related myeloid cancers. Viewing tumor biology in the context of CH also offers a novel perspective for biological discovery in cancer research. Still, the impact of CH is nuanced, and altered molecular pathways are likely to be cancer-specific, warranting further investigation. In our survey of CH and its consequences in solid cancer, we advance the CPTAC mission of studying the molecular basis of cancer by focussing on somatic alterations in the immune cells, rather than the tumor. We envision a future where CH and immuno-genomics can be integrated with molecular tumor diagnostics to unlock the full potential of personalized cancer care.

## Methods

### Resource Availability

For CH variant filtering scripts, see Vlasschaert *et al*.^26^

Additional supplemental data and scripts used to conduct this study will be released upon publication of the manuscript.

Inquiries regarding data and code access should be directed to the corresponding authors.

### Study Approval & Data Access

Human research protocol was approved by the Queen’s University Health Sciences Research Ethics Board (PATH-29-22). Informed consent was obtained by the participating CPTAC and TCGA partner projects, with access to controlled raw genomic and transcriptomic data granted by the NCI data access committee (dbGaP Project #33927) via the NIH NCI genomic data commons portal. Additional patient and sample metadata was collected from several CPTAC and TCGA marker papers.^34,51–64^

### Sample Patient Information

This study was conducted using cancer patient data available from the CPTAC and TCGA cohorts (Table S1). The cancers represented in CPTAC (n=1,550) include breast cancer (BRCA; n=130), clear cell renal cell carcinoma (ccRCC; n=214), colorectal adenocarcinoma (COAD; n=106), glioblastoma multiforme (GBM; n=104), head and neck squamous cell carcinoma (HNSC; n=110), lung adenocarcinoma (LUAD; n=229), lung squamous cell carcinoma (LUSC; n=108), high grade serous ovarian cancer (OV; n=90), pancreatic ductal adenocarcinoma (PDAC; n=163), and uterine corpus endometrial carcinoma (UCEC; n=240), as well as a small number of other cancers from various sites (Other; n=56). TCGA (n=8,927) included 32 different solid tumor types. All patients with available peripheral blood whole exome sequencing (WES) were included in CH variant calling. Only primary, treatment-naïve tumor samples in patients with metadata for all relevant covariates were included in transcriptomic, proteomic, and phosphoproteomic analyses.

### Genomic Data Processing & Clonal Hematopoiesis Variant Calling

Peripheral blood and tumor WES data aligned to the GRCh38.d1.vd1 reference genome was acquired from the CPTAC project and was processed via the Genome Analysis Toolkit (GATK)-Mutect2 v4.2.4.0.0 algorithm to identify potential CH driver mutations.^65^ Depth of coverage data in CH driver genes was collected using GATK-DepthOfCoverage v4.2.4.0.0^65^ There are known challenges in variant calling related to a false duplication of *U2AF1* that precluded the use of GATK-Mutect2 for variant calling in this gene.^66^ To circumvent this, a custom pileup region script was applied as in Vlasschaert *et al*.^26^

Functional annotation of variants was performed with ANNOVAR using the Ensembl database.^67^ The annotated variants were then subject to R-based filtering scripts and manual review as adapted from Vlasschaert *et al*.^26^ to curate the final list of CH driver mutations in the study cohort. For sequencing-based filtering, we required depth of coverage ≥ 20x, alternative allele reads ≥5, and paired-end forward and reverse reads (F1R2/F2R1) ≥1. Insertion/deletion variants occurring at homopolymer sites ≥3 base pairs long were removed. Remaining variants were filtered against a curated list of CH driver mutations. Consistent with standard definitions of CH, VAF ≥2% was required. To eliminate germline variants, variants that had a binomial p>0.01 of an equal number of normal and alternative allele reads (i.e. 50% VAF). Any remaining recurring artefacts were removed if they appeared more frequently than established CH hotspots like *DNMT3A* R882H. During CH calling, three patients exhibited JAK2 V617F mutations with VAF >50% (C3N-03021; TCGA-26-5135, TCGA-DJ-A2Q2). These clones may represent a CH mutation with co-occurring loss of heterozygosity at the same locus, but due to the potential of polycythemia vera in these cases, these patients were excluded from subsequent analyses.

*KMT2C* (formerly *MLL3*) suffers from collapsed duplications in the reference genome, creating challenges for alignment algorithms to appropriately map reads to their origin in the genome.^68^ As a result of this, true variant calls in *KMT2C* are obscured by a high proportion of false positive variants that arise from elsewhere in the genome. Bam files were manually inspected at the *KMT2C* locus using Integrated Genomics Viewer v2.16.1 to identify exons impacted by poor mapping quality – indicative of the collapsed duplication.^69^ Exons 7, 8, 14-20, and 23-25 were found to have evidence of non-specific genome mapping, and were therefore blacklisted due to poor confidence in variant calling. The remaining exons were retained for variant filtering as described above.

Variant filtering and curation to identify CH-Tum was conducted in all tumor samples from patients with CH mutations found in their peripheral blood sample. Somatic variants in each tumor sample were parsed for the same CH mutation(s) that presented in the peripheral blood. Mutations that presented in both the tumor and blood with VAF ≥ 40% and/or a tumor to blood VAF ratio ≥ 2, representing likely germline variants and tumor variants, respectively, were identified as false positive CH calls and removed from the final list of CH mutations.

Oncoplots representing CH and CH-Tum mutations were created for the 15 most frequently mutated genes using cBioPortal Oncoprinter v6.0.5.^70,71^

### Survival Analysis

Differences in overall survival based on CH, CH-Tum, CH-PB, and *DNMT3A* and *TET2* status were evaluated using univariate and multivariate Cox proportional hazard models provided by the survival v3.5-7 R package, with Kaplan-Meier curves illustrated with the survminer v0.4.9 R package. The modelled covariates in survival analysis included patient age (years), sex, smoking (smoker versus non-smoker), and disease stage (local versus metastatic). Cancer type was controlled for in pan-cancer analyses. CIBERSORTx immune infiltrate scoring (described below) was also included as a covariate where stated. In the mesenchymal glioblastoma subset, *NF1* mutation status was also included as a covariate. Modelling of last known tumor status at follow up excluded patients with metastatic disease. Significant results were indicated by 95% confidence intervals that did not overlap with HR = 1 and p < 0.05.

### RNA-seq Data Processing

Aligned bulk tumor RNA-seq bam files were de-aligned into raw, unstranded, paired-end fastq files with samtools v1.17.^72^ Quality filtering and adapter trimming of fastq files was performed using fastp v0.23.1, where sequencing reads with base Phred score ≥ 15, and ≤ 40% bases unqualified were retained in the fastq files.^73^ Reads in the filtered paired-end fastq files were pseudo-aligned and transcript expression was quantified using kallisto v0.46.1 based on release 109 of the Ensembl cDNA reference for GRCh38.^74^ Reports were collected in MultiQC v1.13 for review of sequencing and quantification quality.^75^

### Differential Gene Expression Analysis

Differential gene expression analysis was performed using DESeq2 v1.26.0 to identify variations in the transcriptional landscape of the TME based on CH, CH-Tum, CH-PB, and *DNMT3A* and *TET2* status.^76^ Transcript quantifications generated by kallisto were aggregated to the gene level and imported into DESeq2 with tximport v1.28.0.^77^ Genes with low abundance were filtered out if they showed expression counts < 10 in *n* samples, where *n* represents the number of samples with CH. Non-coding genes were also removed from the analysis. Analyses was conducted with the following covariates: PAM50 subtype for BRCA, microsatellite instability status for COAD, and *NF1* mutation status for mesenchymal GBM, as well as patient sex where appropriate. HPV-positive HNSC and non-clear cell renal cell carcinomas were excluded. Cancer type was controlled for in pan-cancer analyses. Independent hypothesis weighting methods from the IHW v1.14.0 package were used to derive multiple testing adjusted p values.^78^ Adjusted p < 0.05 was required for a gene to be considered differentially expressed. Generated log fold change values were subject to ashr v2.2-63 effect size shrinkage for the purposes of ranking and downstream analysis via gene set enrichment analysis (GSEA).^79^ Volcano plots displaying DGE were generated using the EnhancedVolcano v1.20.0 R package. Venn diagrams of differentially expressed genes in CPTAC and TCGA were created using the ggvenn v0.1.10 R package. Overlapping differentially expressed genes in CH-Tum were subject to functional profiling of Gene Ontology (GO) terms using g:Profiler g:GOSt with default settings.^80^

### Deconvolution of Immune Cell Subpopulations

Normalized RNA expression counts were obtained from DESeq2 and loaded into the CIBERSORTx cell fractions module for deconvolution. Deconvolution was based on the LM22 immune gene signature matrix with B mode batch correction and disabled quantile normalization, and the analysis was run in absolute mode with 100 permutations.

Abundances of certain LM22 populations were aggregated to create derivative cell populations as listed below:

Monocyte/Macrophages = Monocytes + M0 Macrophages + M1 Macrophages + M2 Macrophages

Macrophages = M0 Macrophages + M1 Macrophages + M2 Macrophages

B Cells = Naïve B Cells + Memory B Cells + Plasma Cells

T Cells = CD8+ T Cells + Naïve CD4+ T Cells + Resting Memory CD4+ T Cells + Activated Memory CD4+ T Cells + Follicular Helper T Cells + Regulatory T Cells + γδ T Cells

CD4+ T Cells = Naïve CD4+ T Cells + Resting Memory CD4+ T Cells + Activated Memory CD4+ T Cells + Follicular Helper T Cells + Regulatory T Cells

NK Cells = Resting NK Cells + Activated NK Cells

Dendritic Cells = Resting Dendritic Cells + Activated Dendritic Cells

Absolute scoring of cell type abundances by CIBERSORTx is described elsewhere,^81,82^ and represents the median expression of cell type-specific genes relative to median expression of all genes in the sample. Samples with deconvolution p ≥ 0.05 were deemed unreliable for further analysis. Cell type abundances were compared for CH, CH-Tum, CH-PB, and *DNMT3A* and *TET2* status with linear regression using a Tweedie distribution, adjusting for the same covariates used in differential expression analysis described above.

### Differential Protein & Phosphoprotein Abundance Analysis

Proteomics and phosphoproteomics data pre-processing is extensively described in Li *et al*.^51^ Differential protein and phosphoprotein analysis for CH, CH-Tum, CH-PB, and *DNMT3A* and *TET2* status was conducted using limma v3.58.1 with the same covariates described above, with adjusted p values derived using the Benjamini-Hochburg correction.^88^ Proteins or phosphoproteins with low abundance were filtered out if they held an NA value in *n* samples, where *n* represents the number of samples with CH. Adjusted p < 0.05 was required for a protein to be considered differentially expressed.

### Gene Set Enrichment Analysis

Enrichment of biologically relevant pathways at the transcriptomic and proteomic level was evaluated with GSEA v4.3.2.^29,30,89^ Analysis was performed using GSEAPreranked with 1000 permutations and weighted enrichment statistics. Rank scores of features input to GSEA was calculated as -log(p)*sign(log[FC]). All comparisons made were subject to GSEA for the HALLMARK gene sets and three gene sets predictive of response to anti-PD-1 ICI therapies.^35,36,85^ In GBM, a glioma stem cell signature (split between upregulated and downregulated genes) was tested in CH-Tum.^35,36,85^ Results for CH-Tum in mesenchymal GBM were additionally subject to GSEA using the extended EnrichmentMap gene sets v2024-01-01 available from the Bader lab and GBM-derived immune cell phenotype signatures.^39^ FDR < 0.1 was required for a gene set to be considered significantly enriched.

GSEA output from the EnrichmentMap gene sets was loaded into Cytoscape v3.10.1 using EnrichmentMap v3.3.6.^86,87^ Gene sets were clustered using clusterMaker2 v2.3.4 and AutoAnnotate v1.4.1, with the 15 largest clusters retained in the figure. Cluster labels were manually generated to summarize the included gene sets.

### Kinase-Substrate Enrichment Analysis

The fold change and p values for each phosphoprotein calculated by limma was input into the KSEA App v1.0 to infer relative activity of kinases.^88^ Kinase-substrate enrichment analysis (KSEA) was run using the PhosphoSitePlus and NetworKIN databases following the default app settings.^84,97,98^ Kinases were considered to be significantly enriched with an FDR < 0.1. Positively enriched kinases were then passed into g:Profiler g:GOSt for functional profiling, using the KEGG and Reactome pathway databases and default web settings.^80,91,92^

### Statistical Analysis

All statistical analyses were conducted using R v4.3.1. Figures were produced using ggplot2 v3.5.0, ggridges v0.5.6, and ggbeeswarm v0.7.2, along with various other R packages as stated above. Schematic Figure 1A was created with BioRender.com. Statistical tests employed for analysis include the Wilcoxon rank-sum test, Spearman’s rank-order correlation, and generalized linear models incorporating binomial and tweedie distributions. The wec v0.4-1 R package was used to generate weighted mean contrasts for logistic regression modelling of CH and CH-Tum ORs by cancer type.^93^ The polr function of the MASS v7.3-60.0.1 R package was used to conduct ordered logistic regression for cancer stage analyses. The glmmTMB v1.1.9 was used to model CIBERSORTx fractions using linear regression with the Tweedie distribution.

## Data Availability

Supplemental data and scripts used to conduct this study will be released via Zenodo upon publication of the manuscript. Inquiries regarding data and code access should be directed to the corresponding authors.

## Acknowledgements

M.M.B. was supported by a CGS-M scholarship from the Canadian Institutes for Health Research.

C.V. was supported by a CGS-D scholarship from the Canadian Institutes for Health Research.

A.G.B. was supported by the National Institutes of Health (Grant No. DP5-OD029586), the Burroughs Wellcome Foundation Career Award for Medical Scientists, and the Pew-Stewart Scholar for Cancer Research award.

R.J.V. was supported by the Leukemia & Lymphoma Society of Canada with Canadian Institutes for Health Research, a Brain Tumour Foundation of Canada Elevation Grant, and the Princess Margaret Cancer Foundation.

M.J.R. is supported by grant funding from the Canadian Institutes for Health Research (Project Grant No. 202010PJT-451137) and Ontario Institute for Cancer Research (Project No. CPTRG-056).

We would like to acknowledge the CPTAC and TCGA programs for providing the data used in this study and the Centre for Advanced Computing at Queen’s University for providing the high-performance computing resources and support to conduct study analyses. We also thank Mark D. Minden, Stanley W.K. Ng, and Andy G.X. Zeng for their thoughtful comments and suggestions.

## Author Contributions

Conceptualization: All authors

Formal Analysis: M.M.B., R.J.V., M.J.R.

Funding Acquisition: R.J.V., M.J.R.

Methodology: All authors

Supervision: R.J.V., M.J.R.

Visualization: M.M.B.

Writing – Original Draft: M.M.B.

Writing – Review & Editing: All authors

---

**Figure S1:**
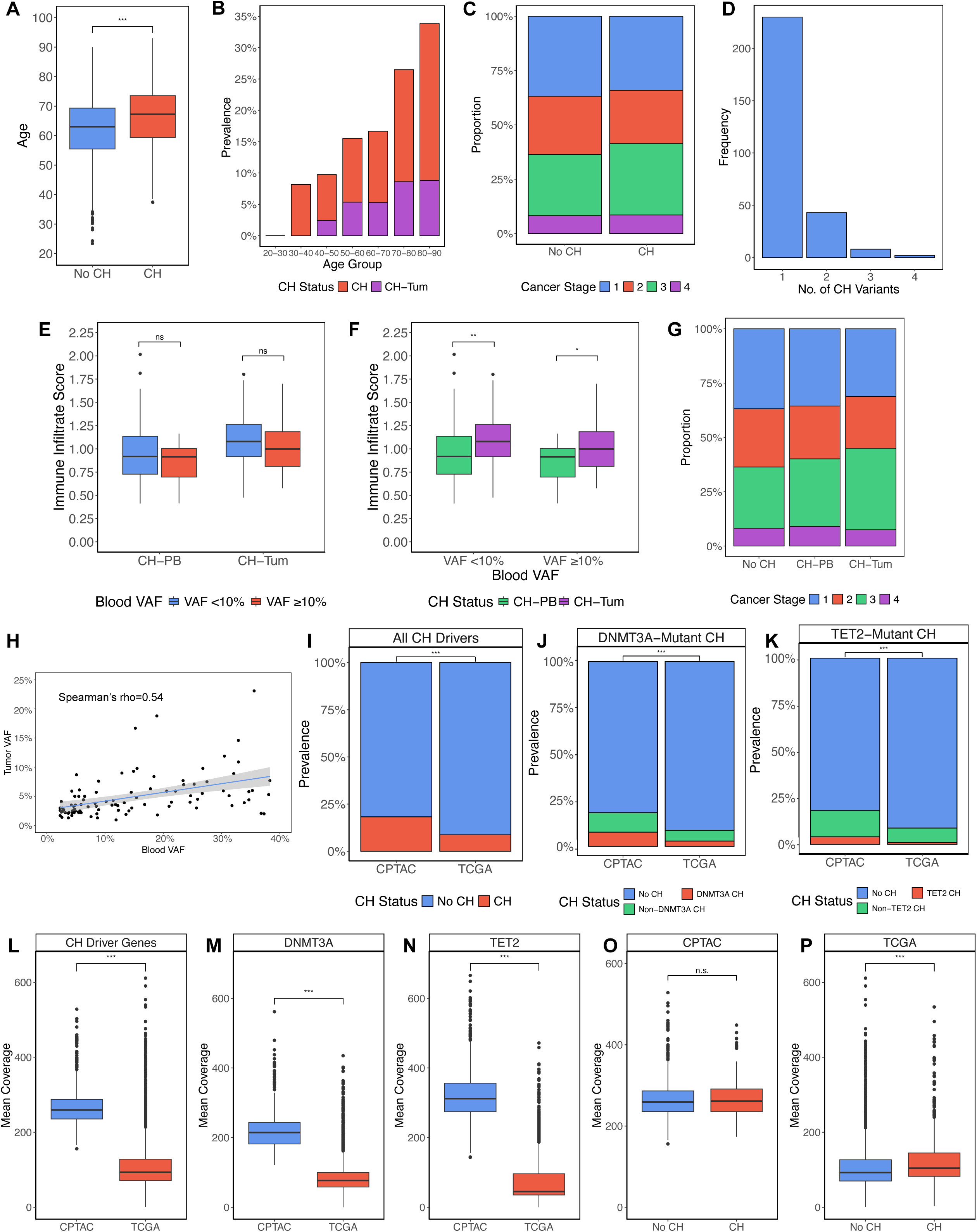
Survey of the characteristics of CH and CH-Tum in treatment-naïve solid cancer patients, related to Figure 1. (A) Patient age for those with and without CH. (B) Prevalence of CH and CH-Tum by age decade. (C) Tumor stage at diagnosis by CH status. (D) Number of CH driver mutations in patients with CH. (E) Immune infiltrate scoring based on peripheral blood VAF in CH-PB and CH-Tum. (F) Immune infiltrate scoring based on CH-Tum status in large and small CH clones. (G) Tumor stage at diagnosis by CH-Tum status. (H) Correlation of mutation VAF for CH mutations in peripheral blood and tumour samples. (I-K) CH (I), DNMT3A-mutant CH (J), TET2-mutant CH(K) prevalence in CPTAC and TCGA cohorts. (L-N) Average depth of coverage in CPTAC and TCGA cohorts across all CH driver genes (L), DNMT3A (M), and TET2 (N). (O-P) Average depth of coverage for patients with and without CH for CPTAC (O) and TCGA (P).

**Figure S2:**
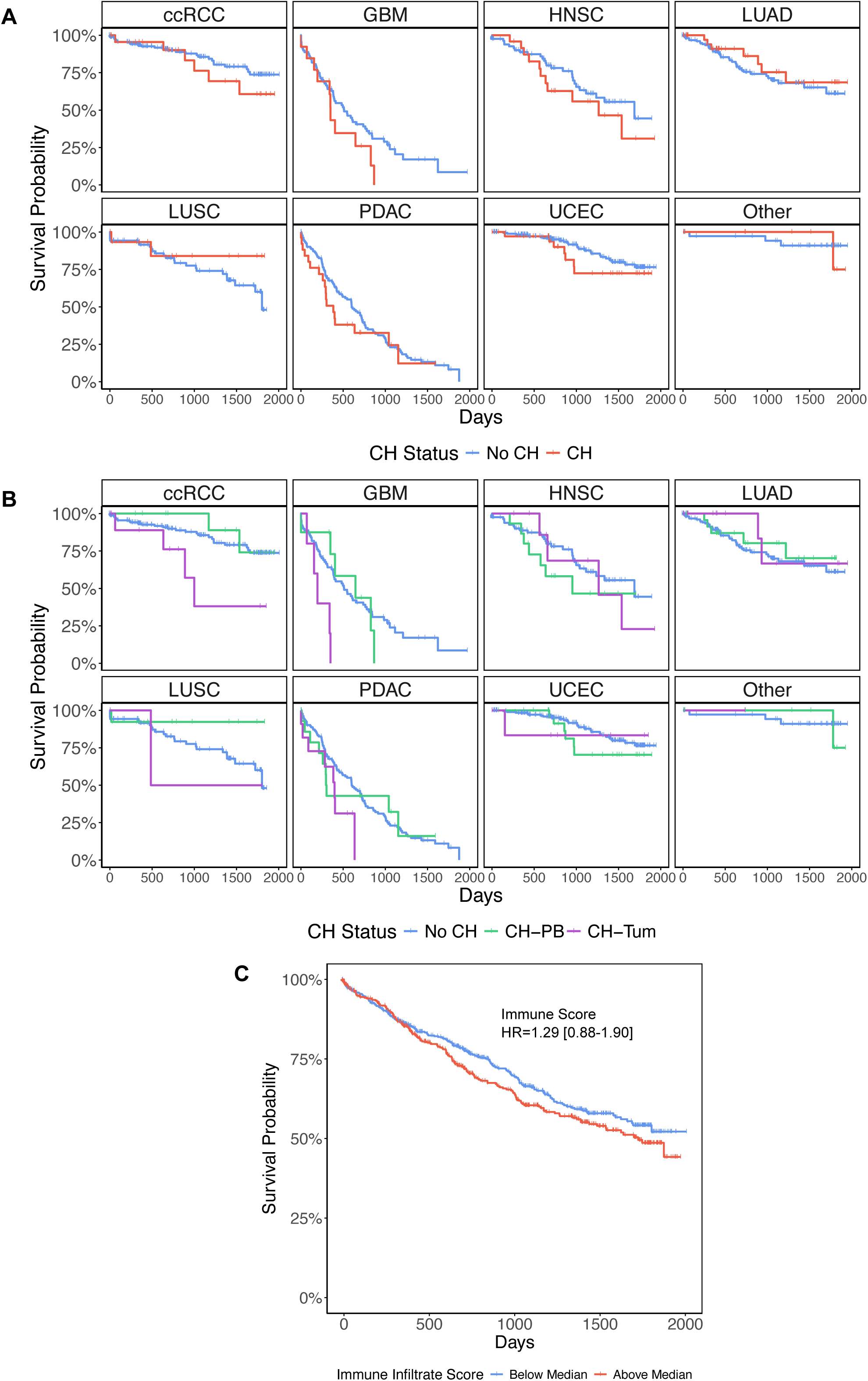
Clinical associations of CH and CH-Tum in solid cancer patients, related to Figure 2. (A) Kaplan-Meier plot comparing overall survival in those with and without CH, stratified by cancer type. (B) Kaplan-Meier plot comparing overall survival in those with CH-Tum, CH-PB, and no CH, stratified by cancer type. (C) Kaplan-Meier plot comparing overall survival based on immune infiltrate scoring.

**Figure S3:**
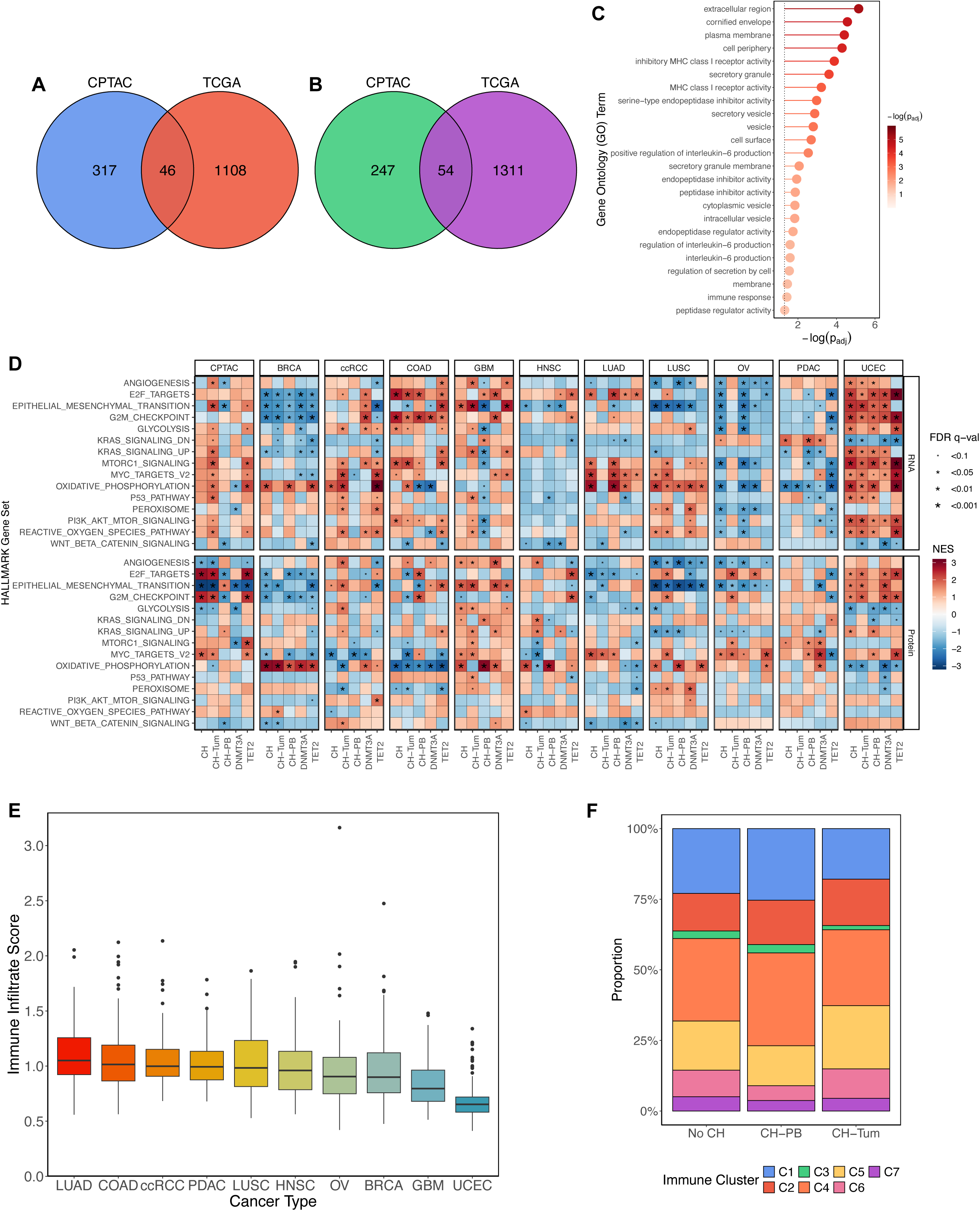
Transcriptomic and proteomic associations of CH in the TME, related to Figure 3. (A-B) Venn diagrams of differentially expressed genes shared by CPTAC and TCGA for CH (A) and CH-Tum (B) (C) Significantly enriched GO terms amongst the differentially expressed genes in CH-Tum common in CPTAC and TCGA (D) Heatmap displaying RNA and protein GSEA of select tumor-associated HALLMARK gene sets by CH status. (E) Immune infiltrate scoring in CPTAC by cancer type. (F) Immune cluster assignment for tumors based on CH status.

**Figure S4:**
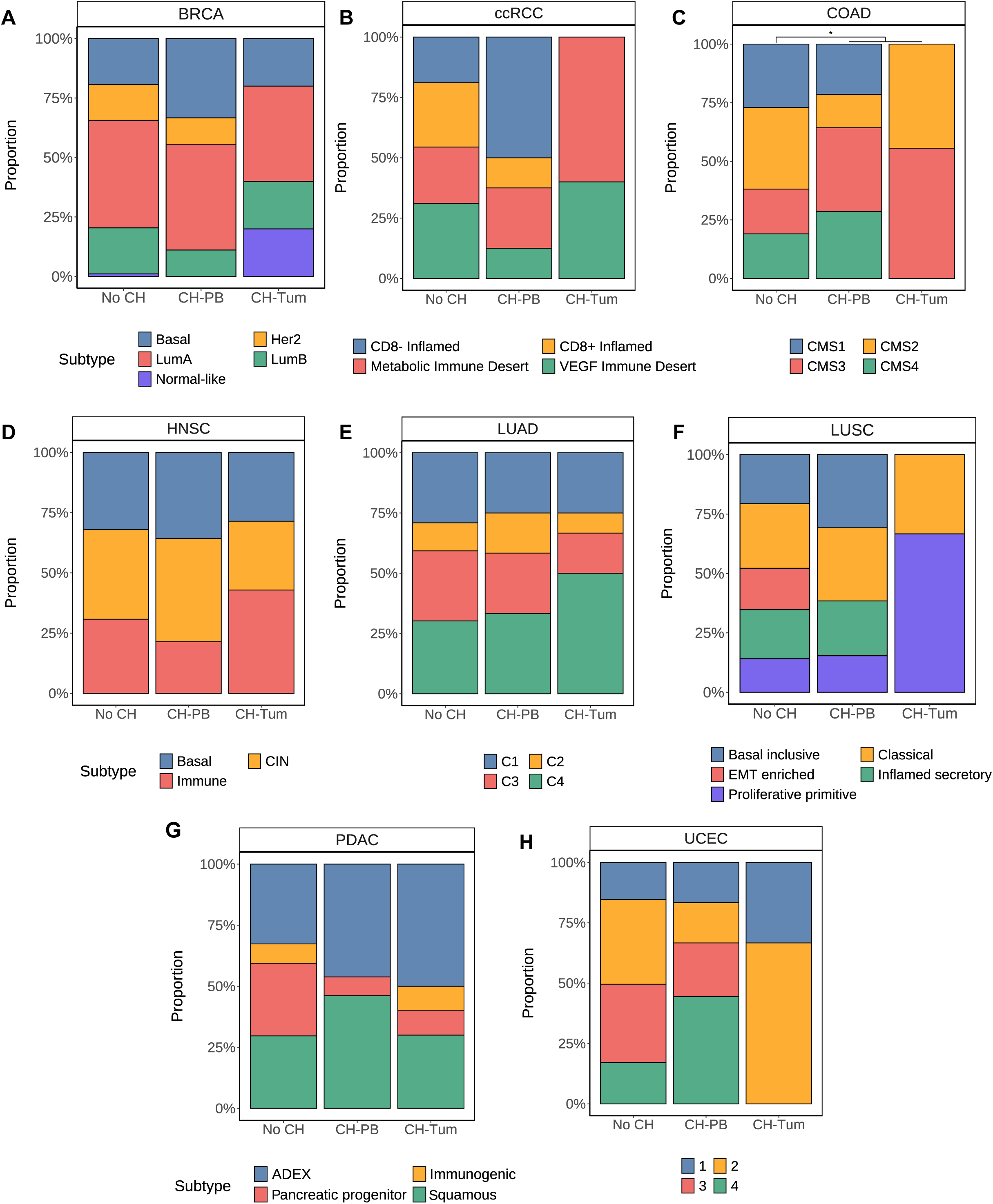
Tumor molecular subtyping associations with CH. (A) BRCA PAM-50 subtypes by CH status. (B) ccRCC molecular subtypes by CH status. (C) COAD CMS subtypes by CH status. (D) HNSC molecular subtypes by CH status. (E) LUAD molecular subtypes by CH status. (F) LUSC molecular subtypes by CH status. (G) PDAC molecular subtypes by CH status. (H) UCEC molecular subtypes by CH status.

